# Enhancing family planning services in northern Ghana: a landscape assessment to develop strategic interventions using an academic partnership approach

**DOI:** 10.64898/2026.01.22.26344657

**Authors:** Sasha Hernandez, Cibel Quinteros Baumgart, Kate Tolleson, Hawa Malechi, Athanasius Ayete Labi, Sunday Imogie, Rayza Sison, Marie A. Brault, Ana Maria Simono Charadan

## Abstract

Northern Ghana faces a significant shortage of family planning (FP) services, leading to high rates of unplanned pregnancies and unsafe abortions that contribute to regional maternal mortality. Through the Academic Model Providing Access to Healthcare (AMPATH) Ghana, a partnership between Tamale Teaching Hospital (TTH), the University for Development Studies School of Medicine (UDS-SoM), and New York University Grossman School of Medicine (NYUGSOM), key stakeholders began strengthening regional FP services using the Exploration-Preparation-Implementation-Sustainment (EPIS) framework. We present how an adapted landscape assessment was used during the Exploration and Preparation phase to guide intervention development at TTH, the region’s sole tertiary referral FP clinic.

We adapted the Supply–Enabling Environment–Demand (SEED^TM^) Assessment Guide for Family Planning Programming. The original survey, which included both open-ended and closed-ended questions, was edited and contextualized by stakeholders at TTH, UDS, and NYUGSOM, pilot tested (n=8), and distributed to clinicians at the FP unit (n=24). Responses were analyzed and grouped by key themes. Areas for improvement included staff training, especially youth-friendly delivery, service integration with a focus on cervical cancer screening and abortion care, and barriers to access for underserved groups including youth, adolescents, and single men. Stakeholder review of survey results highlighted three priorities: in-service trainings, integration of cervical cancer screening, and youth-centered counseling training.

The landscape assessment provided clear direction for strengthening FP services at TTH and set the foundation for AMPATH Ghana’s next steps. The priorities identified offer a focused roadmap for improving training, service integration, and access. As the team advances into the Implementation and Sustainment phases of the EPIS framework, these findings will guide coordinated action and demonstrate the value of implementation science in building effective and equitable interventions within global health partnerships.

## Introduction

Despite Ghana’s rapid economic development over the past 30 years, the benefits have not been evenly distributed in the country, with significant socioeconomic disparities between the northern and southern regions (Fig 1) [1]. More than half of Ghana’s poor live in the north, with poverty increasingly concentrated among female-headed households [2]. Youth in northern Ghana are three to five times more likely to never attend school compared to their peers in southern Ghana, and the region has the lowest female literacy rate in the country at 44.3% [3]. Compared to women in southern Ghana, those in northern Ghana have less access to prenatal care and safe abortion services, and face higher rates of unplanned and high-risk pregnancies, contributing to increased maternal morbidity and mortality [4–7]. This highlights the need to expand culturally sensitive contraception and abortion care.

**Fig 1.**
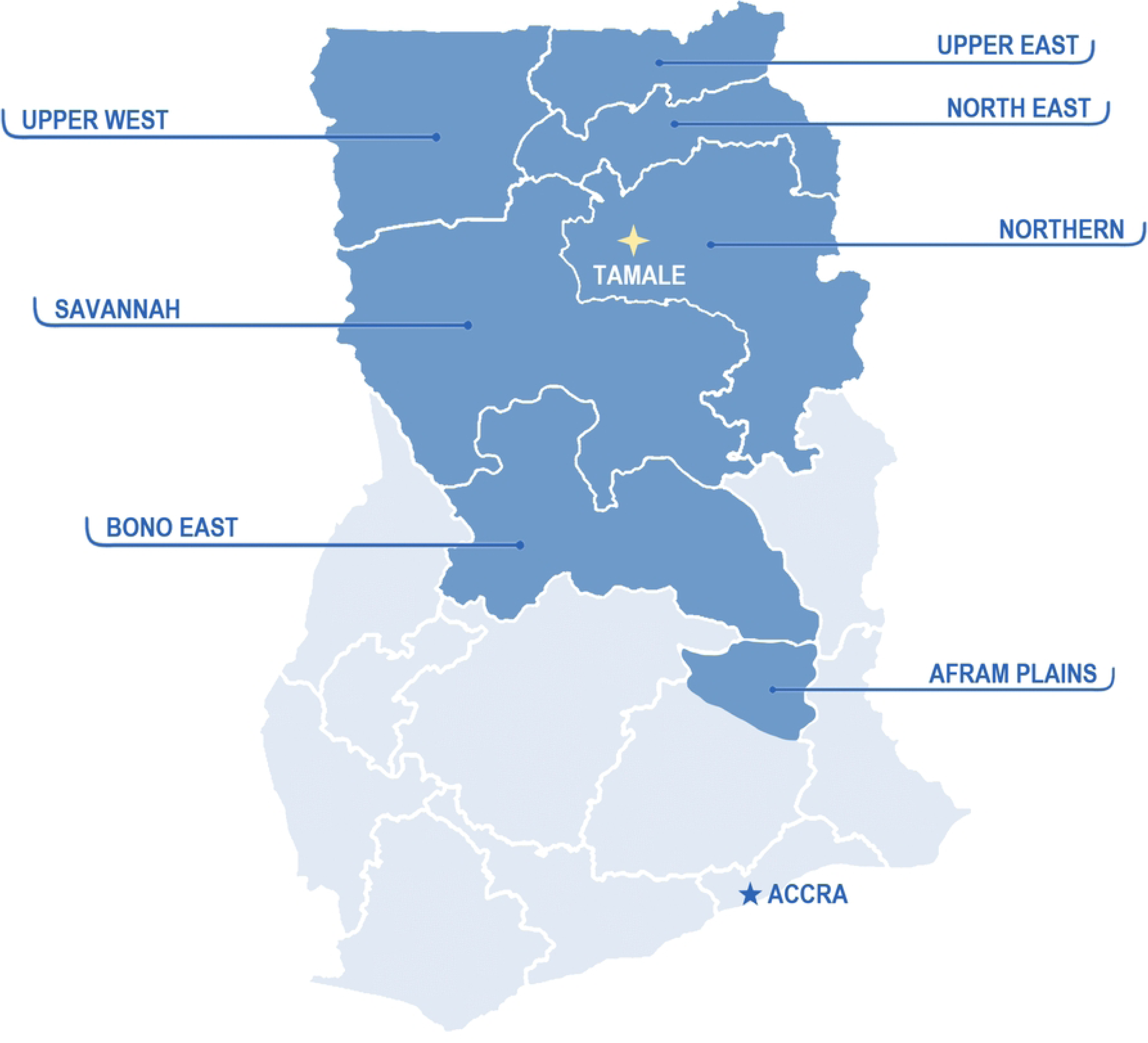
Map of North and South geographic regions of Ghana. Map of Ghana with northern regions highlighted in darker blue and southern regions shown in lighter shading. The location of Tamale and Tamale Teaching Hospital is indicated within the Northern Region.

Multiple health system-level factors, aligned with the World Health Organization (WHO) health system building blocks, influence the expansion, delivery, and uptake of family planning (FP) services in northern Ghana [8]. Within service delivery, challenges include limited integration of FP and abortion-related care, inconsistent availability of services across facilities and communities, and missed opportunities for post-abortion family planning [4, 9–14]. Although both facility- and community-based contraceptive services exist, utilization remains low, suggesting gaps in continuity of care, counseling, and follow-up within routine service delivery. Abortion care remains a lower-priority component within FP service delivery, which may affect access and coordination of care [12, 15]. Constraints related to the health workforce also shape FP uptake, including variability in provider training, limited time for comprehensive counseling, and challenges in addressing client concerns such as side effects, fertility fears, and partner involvement [10, 13, 14, 16]. These workforce-related factors can influence client experiences and trust in FP services. Barriers linked to health financing and access to essential services persist, particularly costs associated with care and geographic distance to facilities, which disproportionately affect rural and low-income populations. Weaknesses in information and service linkages, including referral pathways and follow-up mechanisms, further limit sustained contraceptive use, particularly after abortion or childbirth.

These system-level challenges are especially relevant for youth, first-time mothers, and post-abortion clients, who require tailored, youth-friendly and continuous FP services. For these populations, gaps between contraceptive knowledge and use are shaped not only by individual or social factors but also by health system characteristics, including service organization, provider-client interactions, and availability of supportive care. Addressing these health system dimensions may improve equitable access to FP services and reduce unmet need for contraception in northern Ghana [17–20].

Tamale Teaching Hospital (TTH), the sole tertiary referral and teaching hospital in northern Ghana, houses one of the region’s most comprehensive FP clinics and is well positioned to address existing gaps in reproductive health services especially for these vulnerable populations. Through the Academic Model Providing Access to Healthcare (AMPATH) Ghana, a global health partnership between TTH, the University for Development Studies School of Medicine (UDS-SoM), and New York University Grossman School of Medicine (NYUGSOM) dedicated to advancing health equity, key stakeholders from all three institutions initiated a collaborative effort in 2022 to strengthen FP service delivery in northern Ghana [21,22]. AMPATH Ghana’s Sexual and Reproductive Health (SRH) goals are aligned with the WHO and the Ministry of Health of Ghana who have both emphasized the need to improve FP services to enhance the well-being of women and girls and reduce maternal mortality rates in northern Ghana [16,23]. Leading this effort is AMPATH Ghana’s SRH Team, a key group of women’s health clinicians, educators, and researchers, which applies the Exploration-Preparation-Implementation-Sustainment (EPIS) framework to guide its work. This approach supports a deeper understanding of the service context, fosters stakeholder engagement, promotes culturally competent, sustainable interventions [24], and ultimately helps facilitate the translation of research findings into practical applications, particularly in complex healthcare settings.

While prior research has documented multiple, interacting barriers to FP access in northern Ghana, these barriers span individual, community, and system levels. Less is known about how healthcare system and clinical-context factors specifically shape FP service delivery and uptake, or which aspects of care delivery require strengthening to support effective intervention design. A clearer understanding of how FP services function within clinical settings is needed to inform targeted, feasible, and equitable health system interventions. In this paper, we describe how AMPATH Ghana’s SRH Team applied the EPIS implementation framework [24] to guide planning for FP service improvements at TTH and ultimately the region of northern Ghana. We present findings from a SEED^TM^ (Strengthening Evidence for Effectiveness and Development)-based landscape assessment that identified health system and clinical-context factors influencing FP service delivery during our landscape assessment, laying the groundwork for intentional and context-responsive intervention development.

## Materials and methods

Our study was guided by an overarching academic partnership approach grounded in the EPIS framework as shown in Fig 2 [24]. This section details the Exploration and Preparation phases, explains how they shaped our work, and describes how findings were prioritized to guide intervention development.

**Fig 2.**
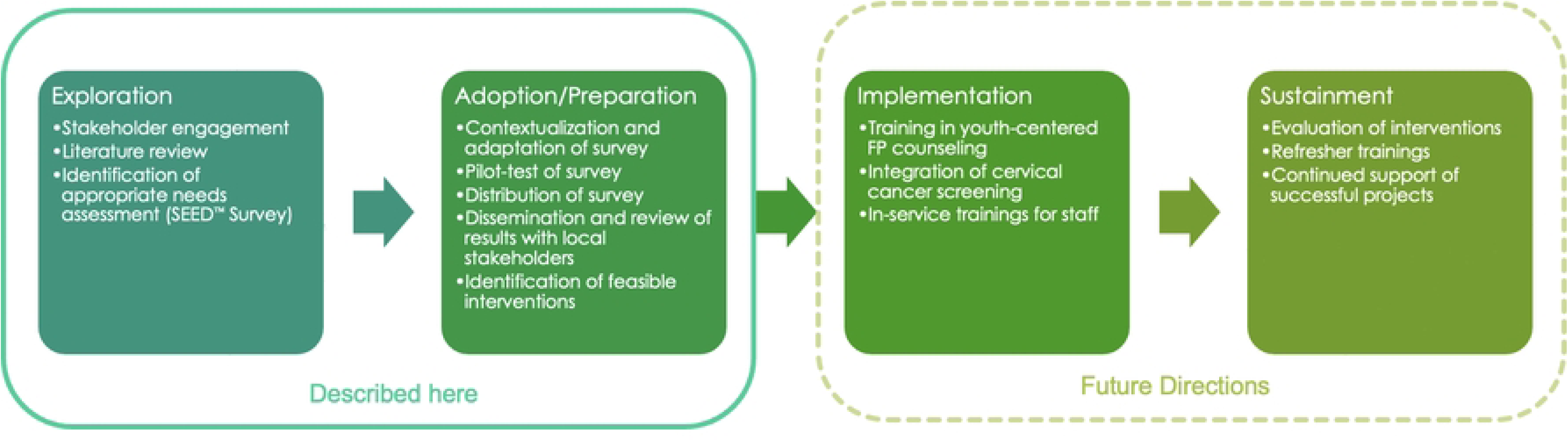
EPIS framework as applied in this project. Overview of the Exploration, Adoption/Preparation, Implementation, and Sustainment phases used to guide intervention development and implementation at Tamale Teaching Hospital.

### Exploration phase

The Exploration phase involved a comprehensive assessment of the needs, resources, and contextual factors relevant to the FP unit at TTH. This phase included three key activities: (1) a review of the literature, (2) stakeholder engagement, and (3) identification of an appropriate landscape assessment tool.

To ground our work in current evidence, we first conducted a literature review to identify best practices and recent interventions in FP service delivery in northern Ghana. Concurrently, and in line with AMPATH Ghana’s standard approach, stakeholder engagement was initiated. Each AMPATH Ghana initiative is co-led by clinical leads from all partner institutions, and a multi-component engagement strategy is employed to ensure alignment with institutional priorities and to secure leadership support. For this project, initial engagement involved formal meetings with the Head of the Department of Obstetrics and Gynecology and the Head of the FP unit at TTH. Their approval was essential to facilitate broader involvement and identify staff members interested in participating in the AMPATH SRH Team.

Following leadership approval, we conducted informal, semi-structured meetings with healthcare providers, administrators, and women who had accessed FP services at TTH. Meetings were conducted separately by stakeholder group and informed by a flexible, topic-guided discussion framework intended to elicit stakeholder perspectives on FP service delivery, access barriers, and opportunities for system strengthening, rather than to generate generalizable research data. These engagements were conducted as part of routine program planning and quality improvement activities and were not designed or undertaken as human subjects research. Accordingly, findings from these meetings are not reported as study results. However, the engagement process played a critical role in stakeholder engagement, trust-building, and informing subsequent implementation planning and decision-making.

One key outcome of this engagement was the selection of an appropriate landscape assessment tool. We identified the SEED^TM^ assessment, developed by EngenderHealth^TM^, as the most suitable option. Designed to help FP organizations comprehensively evaluate program strengths and gaps, SEED^TM^ has been adapted for use in multiple African countries by the RESPOND Project, EngenderHealth^TM^, and the United Nations Population Fund (UNFPA), reinforcing its relevance and adaptability [25–27]. Its holistic and context-sensitive design made it well suited for evaluating FP services at TTH and informing the strategic direction of future initiatives in northern Ghana (Fig 3).

**Fig 3.**
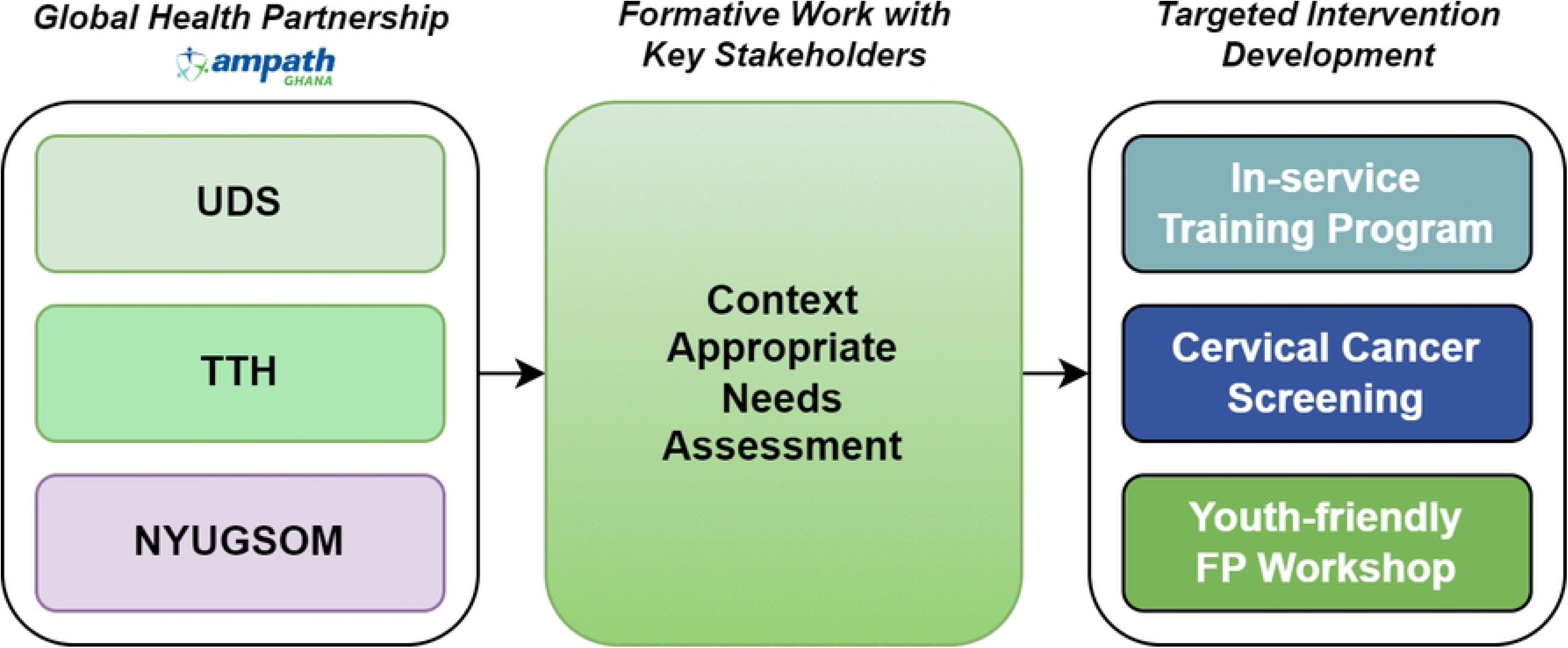
Academic partnership approach to intervention development. Schematic illustrating how the global health partnership between the University for Development Studies (UDS), Tamale Teaching Hospital (TTH), and NYU Grossman School of Medicine (NYUGSOM), through AMPATH Ghana, informed a context-appropriate landscape assessment with key stakeholders, which then guided targeted intervention development.

### Preparation phase

The Preparation phase focused on (1) contextualizing the SEED™ survey for the local setting; (2) pilot-testing the adapted tool to ensure its relevance and clarity; (3) distributing the final version to the target population; and (4) disseminating findings to key stakeholders to identify priority areas for intervention. The SEED™ Assessment Guide was first tailored to the specific needs of the TTH FP unit. Based on prior knowledge of the services offered and cultural insights from local FP providers, we modified the guide by removing questions that were not applicable or outside the scope of the site, adding questions about the roles and FP experience of participants to capture demographic data on FP providers, and reformatting or breaking down some questions to facilitate ease of use in an anonymous online survey with multiple-choice questions and opportunities for free-text responses (S1 Table). The adapted survey was reviewed by multiple Ghanaian FP-trained Obstetrician and Gynecologists to ensure the language and content were appropriate for the cultural context of northern Ghana.

To further ensure the SEED™ survey was appropriate for the local context, we conducted a pilot test with a small cohort (n=8) comprised of staff from the TTH FP unit. The purpose of the pilot was to assess the clarity of the questions, cultural relevance of the content, and overall usability of the tool. Participants were asked to complete the survey and provide structured feedback on language, phrasing, and the comprehensiveness of the items. Their input highlighted minor areas for revision, including adjustments to terminology and question framing to better align with local norms and service delivery realities. This feedback was incorporated into a revised version of the tool, enhancing its user-friendliness and ensuring its effectiveness in capturing meaningful, context-specific data.

#### Data collection

Once the survey was finalized, we distributed it to nurses, midwives, and doctors at TTH. We used targeted recruitment strategies by sharing a link to the online survey via multiple TTH departmental platforms. After reviewing an information sheet, an interested participant was able to proceed to the electronic survey if they were over 18 years old, a clinical provider at the FP unit of TTH and comfortable reading English, the primary language used in medical practice and training at TTH. Surveys were completed independently and anonymously to promote honest responses. Study data were collected and managed using REDCap electronic data capture tools hosted at NYU; a secure, web-based software platform designed to support data capture for research studies [28,29].

#### Data analysis

We downloaded data from REDCap for aggregation and analysis. Univariate statistics were performed to calculate percentages of answers for multiple choice questions out of the total participants who answered that question, and averages and standard deviations calculated for continuous variables (i.e. years worked at the unit). Free text responses were reviewed by the core SRH team and key themes were extracted by group consensus. Responses were then aggregated by theme and salient quotes selected that best represented the ideas expressed.

#### Prioritization of Findings

Upon completion of the FP Landscape Assessment Survey at TTH, the AMPATH Ghana SRH Team compiled the findings into a comprehensive written summary. This document, which detailed survey responses and highlighted key areas for improvement, was disseminated through departmental communication platforms accessible to all survey participants. This ensured transparency and allowed contributors to review the outcomes of their input.

Following dissemination, the AMPATH Ghana SRH Team convened leadership from TTH’s Obstetrics & Gynecology Department to collectively review the results. During this session, leaders independently rated each identified need and potential intervention based on perceived importance and feasibility. This structured prioritization process enabled the team to identify high-impact, actionable areas that aligned with local capacity and institutional goals, informing the direction of subsequent intervention planning.

### Ethical considerations

This study was conducted in accordance with the ethical standards of the NYU Grossman School of Medicine and local policies at Tamale Teaching Hospital, Department of Obstetrics & Gynaecology. The survey-based study received exemption status from the NYU Grossman School of Medicine IRB (i23-01477) under 45 CFR 46.104(d). TTH Obstetrics & Gynaecology department leadership reviewed IRB exemption from NYU Grossman School of Medicine and deemed it appropriate to distribute the survey throughout their department. The recruitment period for the study started on March 28, 2024, and ended on April 4, 2024. Eligible participants were invited to complete an anonymous online survey through approved channels of department communication (WhatsApp department platforms, snowball recruitment). Participants could choose whether to complete the survey and could stop at any time. Consent was implied by participants’ decision to proceed; no written signatures or verbal consent were obtained per IRB exemption protocols. No minors were enrolled.

## Results

### Overview

A total of 24 respondents from the TTH family planning unit participated in the survey out of 24 eligible participants, with 15 respondents fully completing the survey. Data reported here are percentages calculated from the total respondents for each question. Most respondents identified their role as midwife (50%), followed closely by doctors (45.8%), with one respondent (4.2%) indicating they were a nurse. The years of experience among staff varied from one to 15 years at the unit. The key results of this landscape assessment highlight several areas for improvement in the FP clinic at TTH: 1) staff training needs, particularly in youth-friendly service delivery, 2) need for service integration, and 3) barriers to access and low accessibility for underserved groups (Fig 4). These are further described using direct quotations from study participant responses to free text questions in the survey (Table 1).

**Fig 4.**
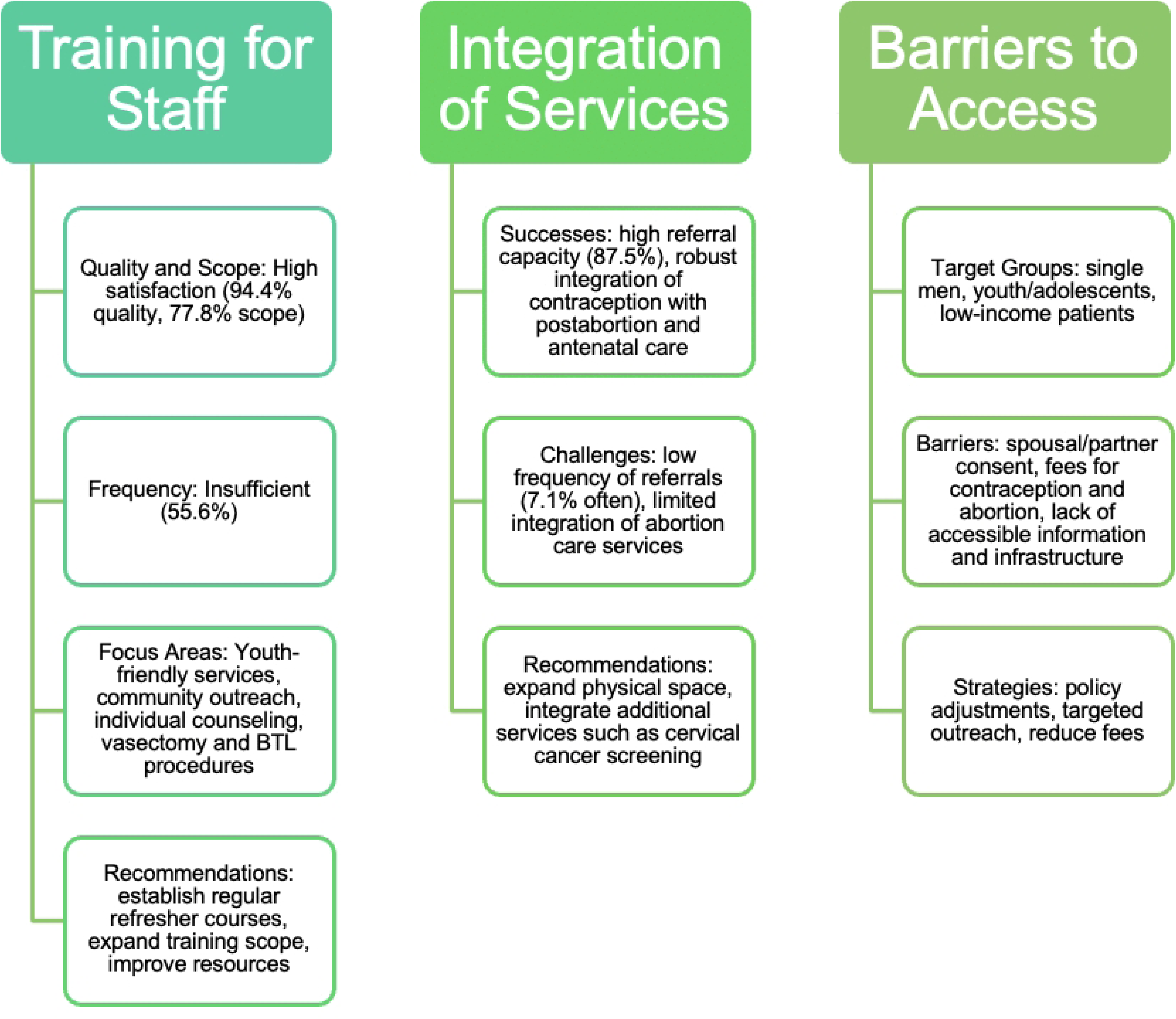
Main findings from TTH FP unit landscape assessment. Overview of key themes identified in the Tamale Teaching Hospital family planning unit landscape assessment, including training needs, service integration gaps, and barriers to access, with corresponding challenges and recommended areas for improvement.

**Table 1.** Participant qualitative survey responses for select sections.

|  |
| --- |
| If FP training is not adequate in regards to quality, please explain why in 1-2 sentences: |

| # | Participant quote |
| --- | --- |
| 1 | “to have yearly refresher studies and to establish side operating room for other methods like BTL and vasectomy. To train people and procure necessary equipment for these” |
| **If FP training is not adequate in regards to scope, please explain why in 1-2 sentences:** |  |
| # | Participant quote |
| 2 | "they are not able to provide services like minilap [mini-laparotomy for female sterilization], vasectomy and others couple with lack of space and privacy for adequate counselling" |
| 3 | "we need residents and house officers to rotate through the [FP] unit to learn." |
| 4 | "we barely performed male sterilization" |
| **If FP training is not adequate in regards to frequency, please explain why in 1-2 sentences:** |  |
| # | Participant quote |
| 5 | “most staff learn on the job without formal training. The GHS always forget(s) about TTH when trainings are organized.” |
| 6 | "In service training is not usually organized by the hospital" |
| 7 | "refresher training not done often" |
| 8 | "Updates in training are not scheduled." |
| 9 | “Frequent training sessions/refreshers have not been conducted and junior doctors' training schedules are not established for adequate lessons.” |
| 10 | "no specialist trained RH present at FP unit at TTH to provide the training" |
| **List 1-3 strengths of the family planning unit at TTH:** |  |
| # | Participant quote |
| 10 | "Well trained and well-motivated staff [and] weekend service" |
| 11 | "Good hygiene, friendly staff, good communication and counseling services" |
| 12 | "Provision of contraceptive methods like the hormonal ones [and] the love for the work by the providers of the services" |
| 13 | "it's able to meet client need despite the few staff strength, able to support peripheral facilities in terms of training, able to deal with deeply inserted implants and missing IUD" |
| 14 | "1. Experienced staff, 2. Easily accessible even on holidays and weekends, 3. Proper documentation" |
| 15 | "improved staff strength, provision of quality service, reduction of clients waiting time" |
| **List 1-3 specific things that you would want to see done to improve the family planning unit at TTH:** |  |
| # | Participant quote |
| 16 | "increase space, retool essential equipment, in-service training for staff" |
| 18 | "more space to allow teaching, sterilization of their own consumables, mini theatre, integrated services with cervical cancer screening, counselling rooms and conducive waiting area" |
| 19 | "More refresher training sessions, availability of several methods of FP, have an effective schedule for junior doctors' training" |
| 20 | "expand the infrastructure, train more people especially those specialized in [reproductive health], provide equipment or material for services, make sure there is unlimited supply of contraceptive methods." |
| 21 | "increase staff strength, allow residents and house officers to rotate within the unit, get a bigger infrastructure or space to be able to perform abortion procedures and theatre for BTL and vasectomy" |
| 22 | "Good infrastructure, in-service training, improved logistics [instruments] ..." |
| 23 | "reduction of clients waiting time, provision of quality service, provision and availability of logistics [instruments], education and training on family planning, improve infrastructure" |
| 24 | "improve infrastructure, constant supply of equipment, education and training on family planning" |
| 25 | "improve infrastructure, education, and training on family planning" |
| 26 | "frequent and adequate supply of equipment, enough space in order to make the family planning unit more effective and adolescent friendly" |
| 27 | "constant supply of equipment, a bigger place for family planning services, adolescent corner [privacy] for adolescents" |

Selected representative quotations from open-ended survey responses provided by family planning unit staff at Tamale Teaching Hospital. Quotes are organized by thematic area, including training adequacy, strengths of the family planning unit, and suggested areas for improvement.
Abbreviations: BTL, bilateral tubal ligation; FP, family planning; GHS, Ghana Health Service; IUD, intrauterine device; RH, reproductive health; TTH, Tamale Teaching Hospital.

In addition to qualitative findings, respondents reported on the availability of clinical services offered through the TTH FP unit. Reported availability of contraception, abortion care, youth and male reproductive health services, and STI care is summarized in Table 2.

**Table 2:** Availability of services at TTH FP unit.

| Service | N (%) (Number of Respondents Reporting Service Availability) | Details |
| --- | --- | --- |
| Contraception | 23 (100%) | Male condom: 22 (95.7%)<br>Female condom: 20 (87%)<br>Oral contraceptive: 22 (95.7%)<br>Patch/ring: 0% |
|  |  | Injectable: 22 (95.7%) |
|  |  | Intrauterine Device (copper): 22 (95.7%) |
|  |  | Intrauterine Device (hormonal): 0 (0%) |
|  |  | Male sterilization: 1 (4.3%) |
|  |  | Female sterilization: 6 (26.1%) |
|  |  | Fertility awareness: 17 (73.9%) |
|  |  | Education on lactational amenorrhea method: 21 (91.3%) |
| **Abortion** | 4 (17.4%) | Medication management 4 (100% of 4) |
|  |  | Dilation and curettage or MVA: 2 (50% of 4) |
|  |  | Dilation and evacuation: 0 (0%) |
| **Male Reproductive Health** | 9 (39.1%) |  |
| **Youth Reproductive Health** | 13 (56.5%) |  |
| **STI Care** | 11 (47.8%) |  |

Reported availability of family planning and related sexual and reproductive health services at the Tamale Teaching Hospital family planning unit, based on survey responses from unit staff. Values are presented as number and percentage of respondents reporting that a given service is available at the unit. For selected services, specific methods offered are listed with associated number and percentage.
Abbreviations: FP, family planning; MVA, manual vacuum aspiration; STI, sexually transmitted infection; TTH, Tamale Teaching Hospital.

### Training for staff

Almost all (94.4%) respondents found the quality of the FP training they received to be sufficient, and 77.8% found the scope adequate. However, only 55.6% indicated their satisfaction with the frequency of training. Responses about the frequency of in-service trainings varied between yearly (56%), every few years (11.1%), at provider request (22.2%) and never (27.8%). Lack of continued training for staff was the most cited constraint to increasing FP services at the TTH family planning unit, cited by 81.3% of respondents. Multiple areas were identified for additional training and are included in Fig 5.

**Fig 5.**
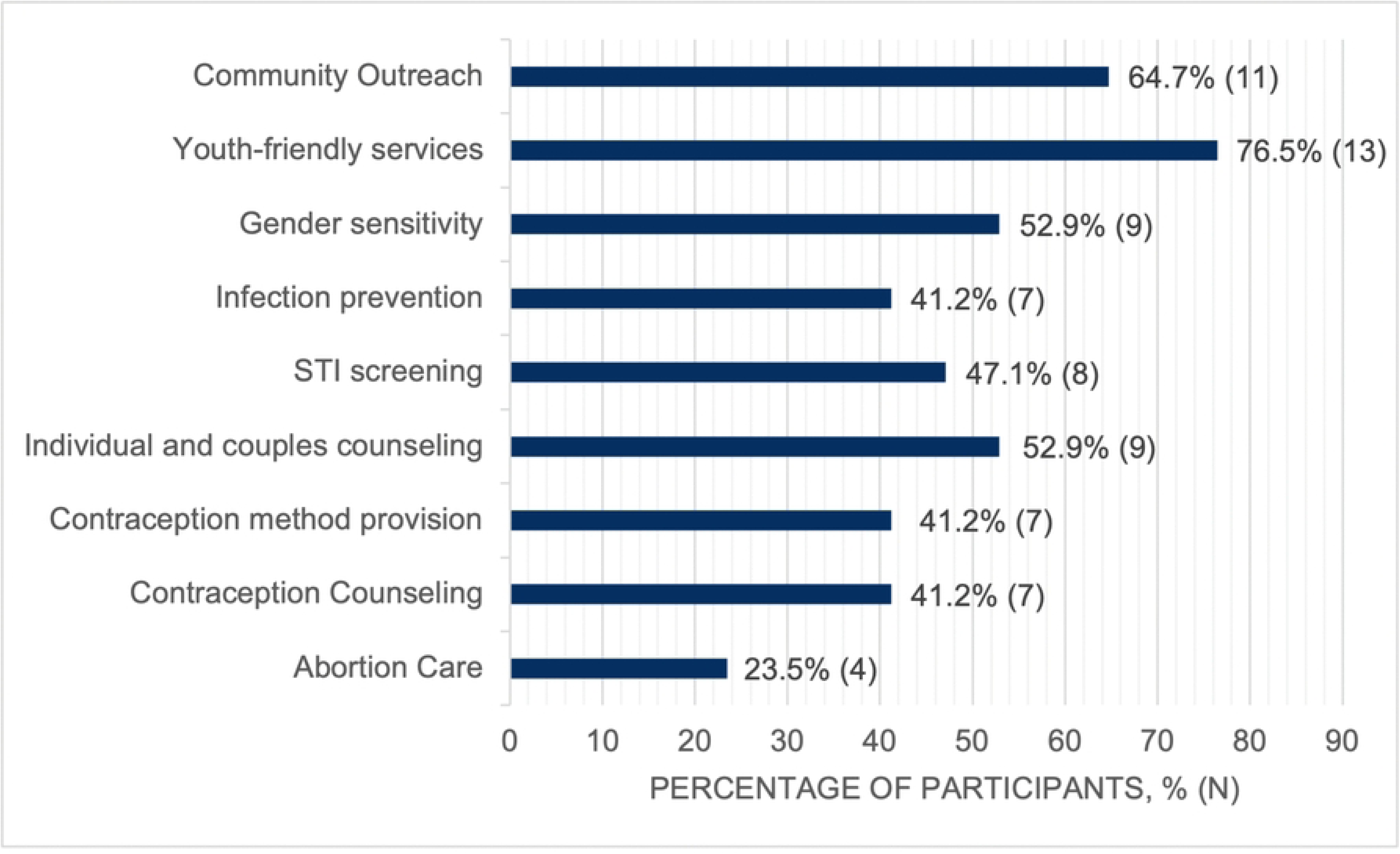
Areas in which participants reported needing additional training. Percentage and number of respondents identifying specific training areas as needing additional training at the Tamale Teaching Hospital family planning unit, based on responses to the landscape assessment survey.

When asked if FP training is adequate in frequency, a doctor with 14+ years of experience raised issues about a lack of formal training and regional disparities in receiving training from larger health organizations: “Most staff learn on the job without formal training. The GHS [Ghana Health Service] always forget(s) about TTH when trainings are organized.”

### Need for further integration of services

The majority of participants reported that contraception care is integrated with postabortion care (93.8% of respondents), postpartum/postnatal care (100% of respondents), antenatal care (93.8% of respondents), and child immunization and well-baby visits (87.5% of respondents). In contrast, abortion care demonstrates comparatively lower integration, with only 37.5% of respondents reporting its integration with primary care, 37.5% reporting integration with antenatal care, 31.3% reporting integration with HIV/AIDS services, and 50% reporting integration with STI services.

When asked to list 1-3 things that they would like to see done to improve the FP unit at TTH, one provider with 2 years of experience wrote: “Integrated services with cervical cancer screening, counselling rooms and conducive waiting area.”

### Barriers to access

Single men and adolescents were described as having the least access to FP services at TTH (Fig 6). While not legally required, 25% of participants report that parental consent is needed for contraception and abortions for adolescents and 37.5% report that spousal consent is required for FP services among married clients. Two respondents suggested that the unit needs to improve its provision of youth-friendly services, identifying a need for more resources, training and a dedicated space for adolescents to promote privacy within the family planning unit. When asked what they believe clients see as their biggest constraints to accessing FP services at the TTH FP unit, 68.8% of respondents reported access, 62.5% reported infrastructure, 43.8% reported lack of information, and 68.8% reported cost. 62.5% of respondents believe that fees influence clients’ choices concerning contraception methods or abortion services.

**Fig 6.**
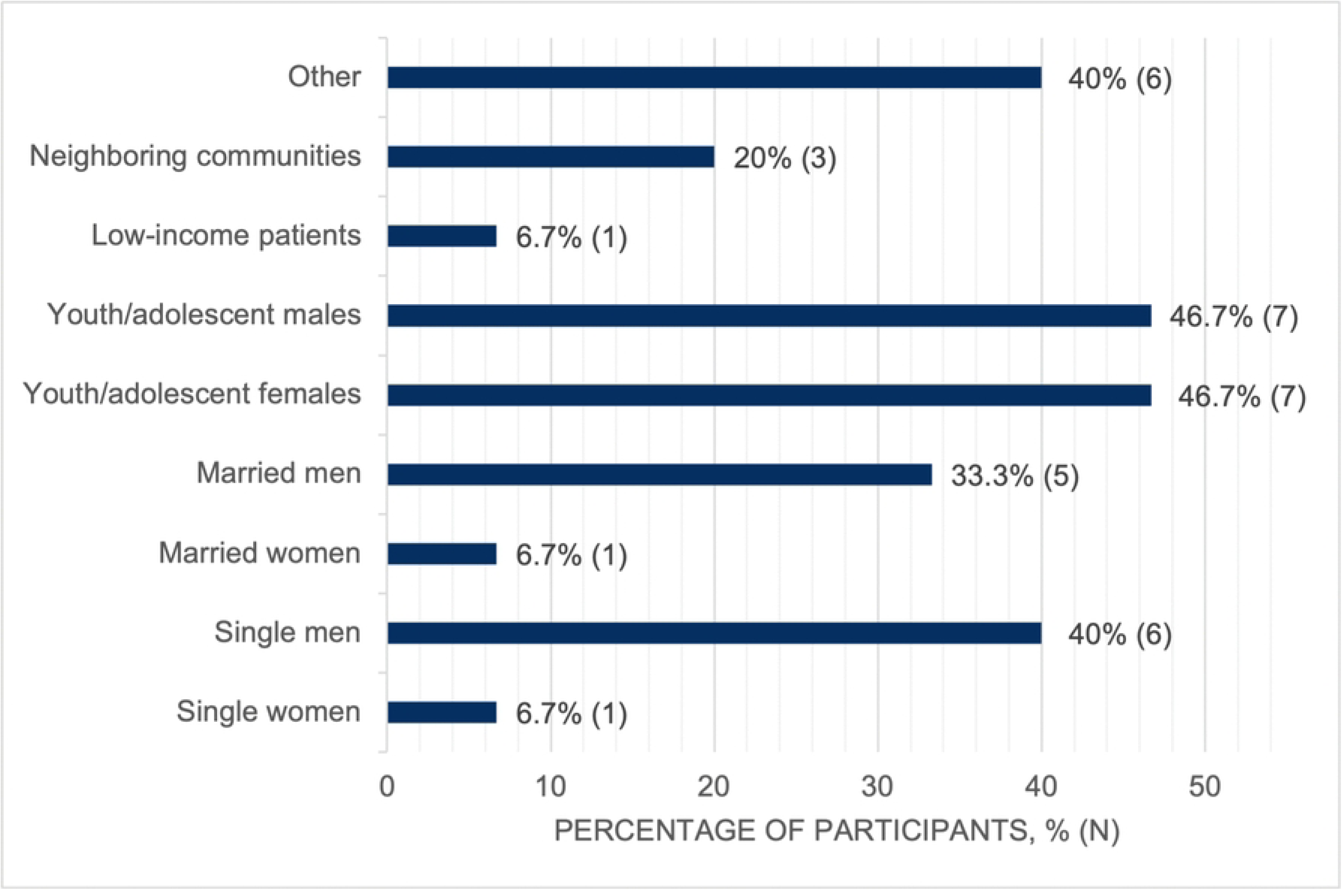
Populations reported to have the least access to the FP unit at TTH. Percentage and number of respondents identifying specific populations as having the least access to services at the Tamale Teaching Hospital family planning unit, based on responses to the landscape assessment survey.

### Data dissemination and priority setting for intervention development

After disseminating findings to the TTH Obstetrics & Gynecology Department, and engaging stakeholders in a prioritization exercise, we identified areas for implementation within a 6–12-month timeframe through AMPATH Ghana. Selected areas were: (1) one to two in-service trainings for staff; (2) integration of cervical cancer screening into FP services; and (3) training on best practices for adolescent-centered family planning counseling.

## Discussion

In summary, we describe how the SEED^TM^ survey was used by the AMPATH Ghana SRH Team to perform a landscape assessment of a FP unit in northern Ghana as part of the Exploration and Preparation phases of the EPIS framework. Our results highlighted several areas for improvement that could be targeted for intervention as part of the Implementation and Sustainment phases of the EPIS framework to ultimately improve FP care and access in the region. We demonstrate that the initial phases of the EPIS framework were successful at identifying areas for intervention using an implementation science lens, and our methods demonstrate the ways in which community and stakeholder engagement are instrumental to our process.

Staff training frequency and quality, as well as the availability of services tailored to vulnerable populations, including adolescents, were identified as priority areas for strengthening through the FP unit landscape assessment at TTH and were subsequently affirmed through stakeholder engagement. These findings align with existing literature from northern Ghana highlighting training and educational initiatives as important strategies for addressing persistent gaps in FP service delivery within the healthcare system. Prior efforts have largely emphasized cultural competency as a core component of patient-centered counseling and have evaluated approaches such as tailored one-to-one counseling and targeted provider training interventions designed to improve care for vulnerable populations [30–34].

Adolescents were identified as one of the populations with the least access to FP services at TTH. Previous research in Ghana has found that adolescents are disproportionately exposed to intrapartum mistreatment [35–37] and recommend provider training that promotes respectful treatment of adolescents [34]. Adolescent sexual and reproductive health has also been a priority of the International Federation of Gynecology and Obstetrics (FIGO) and other organizations on a global scale, with a lack of youth appropriate services being a known contributor to health disparities [38]. Focusing staff training efforts on adolescent-related counseling has been shown to be helpful and a low-resource, high-impact intervention [33].

One of the challenges of achieving true equitable global health partnerships is that priorities are often set by high income country partners. A strength of this work is that we used a multi-phased approach rooted in the EPIS framework with feedback from local stakeholders at every level of the Exploration and Preparation phases to set up a culturally informed foundation for the development and implementation of an intervention. AMPATH Ghana’s SRH team also has leadership equally represented from local and international stakeholders and is built on strong relationships between Ghana and U.S. academic institutions to ensure that priorities are identified by the local community. Additionally, we demonstrated how an objective landscape assessment tool as part of priority setting for a global health partnership can pinpoint essential areas for development, providing a clear intervention roadmap for AMPATH Ghana’s FP initiatives.

Strengths of this assessment include broad dissemination of the survey and participation by all staff members at the target FP unit, with partial or complete responses captured across cadres. The survey generated detailed, practice-relevant information on service organization and delivery that is directly applicable to ongoing service improvement efforts and operational planning for the FP unit and the populations it serves. By collecting data directly from FP unit staff and incorporating feedback from local stakeholders to support shared priority setting, the survey was designed to be culturally appropriate and contextually grounded, thereby enhancing the relevance and potential utility of the findings for informing future interventions.

In addition, the adapted survey instrument and assessment protocol were designed with scalability in mind and may be applicable to other FP units within northern Ghana or adaptable for use in similar clinical settings in other geographic regions. The FP unit at TTH represents one clinical touch point within a broader network of Ghana Health Service facilities delivering FP care across northern Ghana, providing insights that may be relevant to comparable service delivery contexts.

Several considerations should be noted when interpreting these findings. The FP unit staff comprised a relatively small group, and while all staff initiated the survey, 15 of 24 participants completed it in full. Additionally, this assessment focused on a single FP unit within one tertiary institution in northern Ghana. As such, findings are not intended to be globally generalizable but rather to inform local implementation planning and to offer contextually specific insights that may be informative for similar FP service settings within the region.

## Conclusions

This work contributes to the broader discourse on health systems strengthening in underserved regions by demonstrating a structured, stakeholder-driven approach to improving access to high-quality family planning services [9–12,17–19]. The priorities identified through this process align with key WHO health system building blocks, particularly health workforce strengthening, service delivery integration, and the organization of people-centered care for vulnerable populations. Building on the gaps identified during the Exploration and Preparation phases, particularly the need for targeted in-service training, integration of cervical cancer screening, and youth-friendly counseling, future efforts will focus on developing and implementing tangible, contextually relevant interventions. These efforts will be guided by local leadership and delivered through equitable collaboration. Through the Implementation and Sustainment phases of the EPIS framework, these system-level priorities will be operationalized in ways that support locally responsive and sustainable health system strengthening. The upcoming Implementation and Sustainment phases will not only operationalize these priorities but also serve as a model for how implementation science frameworks can support strategic, inclusive intervention development in global health partnerships.

## Data Availability

The survey dataset contains potentially identifying information due to the small sample size and the inclusion of free-text responses therefore, participant-level data cannot be made publicly available. Aggregated data supporting the findings are included in the manuscript. Additional de-identified data may be made available upon reasonable request to the corresponding author, subject to institutional review and execution of an appropriate data use agreement.

## Acknowledgements

We would like to express our sincere gratitude to the Tamale Teaching Hospital family planning unit for their invaluable collaboration and support throughout this work. Their expertise, dedication, and commitment to service delivery were essential to the completion of this assessment and to the broader effort to strengthen family planning services. We are also deeply grateful to the many healthcare providers across northern Ghana who work tirelessly to deliver sexual and reproductive health and family planning services, often under resource-constrained conditions. Their ongoing commitment to patient-centered care is foundational to advancing equitable access to reproductive health services in the region.

## Supporting information

**S1 Table. Adaptation of the SEED^TM^ assessment tool for the TTH FP unit.**

